# Polygenic Embryo Screening: High Approval Despite Substantial Concerns from the U.S. Public

**DOI:** 10.1101/2023.10.13.23297022

**Authors:** Rémy A. Furrer, Dorit Barlevy, Stacey Pereira, Shai Carmi, Todd Lencz, Gabriel Lázaro-Muñoz

## Abstract

Polygenic embryo screening (PES) is a novel – yet commercially available – technology that can compute genetic likelihood estimates of polygenic conditions (e.g., diabetes, depression) and traits (e.g., height, cognitive ability) in embryos. Patients undergoing in vitro fertilization (IVF) can use these polygenic scores to select which embryos to transfer for implantation with the goal of having a child. We conducted a survey of the U.S. public to examine attitudes toward PES encompassing acceptance, interest, potential uses, and concerns (n=1435). Our results indicate 72% public approval for PES, with 82% expressing some interest in using PES if already undergoing IVF. Approval for using PES for embryo selection is notably high for physical health (77%) and psychiatric conditions (72%). In contrast, there is minority approval for embryo selection based on PES for behavioral traits (36%) and physical traits (30%). Nevertheless, concerns about PES leading to false expectations, eugenic practices, and stigma are pronounced (54-55% find them “very” to “extremely” concerning). In a second sample of participants (n=192), presenting concerns at survey onset (vs. end) reduced approval (−28%) by mostly increasing ambivalence (+24%), and only slightly increasing disapproval (+4%). Given its commercial availability, practical limitations, and ethical concerns among physicians, patients, geneticists, bioethicists, and legal scholars, it is notable that there is such high public approval and interest in using PES. Understanding these attitudes is essential for informing policymakers, healthcare professionals, and researchers about the public’s perspectives on this novel biotechnology and debate about the role of medicine in regulating the use of PES.

## Introduction

Polygenic embryo screening (PES), also known as pre-implantation genetic testing for polygenic outcomes (PGT-P), is an emerging biotechnology currently used in the context of in vitro fertilization (IVF) to screen and rank individual embryos for their future genetic likelihood of developing common, complex health conditions. Based on polygenic scores calculated for each available embryo, IVF patients can select which embryo they perceive to be the most desirable to implant for pregnancy. Until recently, pre-implantation genetic testing was limited to monogenic conditions that occur due to single gene mutations (e.g., cystic fibrosis, sickle cell disease)^1,2^. However, with the advent of genome-wide association studies, polygenic scores have been developed for predicting physical health conditions (e.g., coronary heart disease, type 1 diabetes)^3,4^, psychiatric health conditions (e.g., schizophrenia, depression)^5,6^, behavioral traits (e.g., educational attainment, extraversion) ^7,8^ and physical traits (e.g., height, skin color)^9,10^. While studies demonstrate that applying polygenic scores to embryos is practically feasible^11–14^, the technology faces significant limitations due to the correlational nature of polygenic scores, as well as their low predictive power, their variable performance across environments and populations, and the limited genetic diversity between sibling embryos ^15–20^. Beyond these limitations, PES has raised psychosocial concerns among clinicians, patients, genetic counselors, bioethicists, psychologists, and lawyers due to its potential impact on individuals and society at large^9,21–31^. Despite these limitations and concerns, polygenic embryo screening is already commercially available in the U.S. and unregulated as to what types of polygenic outcomes can be screened^32,33^.

Companies offering PES services have presently limited their screening to physical and psychiatric health conditions, but one company has previously offered screening for traits, which suggests it may be offered once again in the future. It is thus of primary importance to understand how the public perceives the use of PES for health conditions and an array of polygenic outcomes. Given the recent emergence of this technology, only two short surveys have published results, and these have been limited to gauging U.S. public approval and interest in PES for specific outcomes^34,35^. In the present survey, we examined 24 conditions and traits - informed by qualitative interviews with US-based REIs and IVF patients^36^. We gauged both approval and interest in varied potential applications of PES. This is also the first survey to examine the public’s views on concerns towards PES. Furthermore, in a second sample, we experimentally investigated the impact of informing the public of potential concerns and the ways this information influences their ratings of approval and interest in PES. Our findings highlight both the importance of comprehensively informing the public about PES and provide insight into how public opinion would shape related policies.

## Methods

### Survey design and analyses

The design of items assessing PES’ potential uses and information types along with potential concerns was informed by qualitative data that was collected from 53 interviews with US-based REIs and IVF patients^36^. Survey participants first received an animated introduction to PES, which briefly explained: IVF, monogenic testing, polygenic testing, as well as multiple different embryo reports that explained and contrasted percentile, average, and absolute risk/chance estimates for several conditions between two embryos. (See supplemental materials for calculation of the risk/chance estimates, which used small to medium plausible effects.) Finally, participants were informed that PES only provides genetic estimates, the environment and chance also play an important role in the development of polygenic conditions, and current polygenic scores are not as accurate for people of non-European ancestries. We included seven comprehension check questions that were programmed to give corrective feedback. As pre-registered, participant data inclusion was restricted to those who answered a minimum of 5/7 questions correctly.

Following the introduction, participants were asked how familiar they were with PES. They were then asked about general approval, how they think they would vote on allowing or disallowing PES, and their perceived risk-to-benefit ratio regarding people accessing this biotechnology. Subsequently, participants were asked to rate their approval of screening embryos for 12 conditions and 12 traits. As pre-registered, we averaged across the twelve traits and twelve conditions and conducted a dependent sample’s t-test to analyze mean differences between traits and conditions. Participants were then presented with a 4×4 matrix and rated whether they approved, disapproved, or did not have an opinion for using PES for the following four purposes that were identified in our qualitative interviews with REIs and IVF patients; information, preparation, embryo selection, and embryo selection based on family history. For each of the purposes, participants rated their approval for four screened outcomes: physical health conditions, psychiatric health conditions, behavioral traits, and physical traits. Responses were summed across outcomes and purposes to run two exploratory repeated measures ANOVAs. Participants also rated their interest in using PES (assuming they were undergoing IVF), their likelihood of undergoing IVF for the purpose of using PES (assuming they wanted a child), and their willingness to pay for PES (assuming the cost of IVF was covered). Participants then rated their level of concern for 13 potential concerns about PES, before finally reporting their demographic information. The survey was created with Qualtrics and distributed by the sampling firm Prolific to nationally representative U.S. participants (stratified on sex, ethnicity, and age). The study was approved by the Baylor College of Medicine Institutional Review Board (protocol H-49262).

As pre-registered, three separate exploratory factor analyses using maximum likelihood and (oblique) Promax rotation for the 12 conditions, 12 traits, and 13 concerns were conducted. The factor analyses for traits and conditions suggested that a 1-factor solution provided the best fit, while a two-factor solution emerged for concerns. The second factor emerging from the concerns was comprised of the following two concerns: inequality due to ethnicity and inequality due to cost.

#### Sample 2 Experiment

In a second sample of participants (N=192), we experimentally randomized whether participants were asked to rate the 13 potential concerns first (at the start of the survey, following the introduction) or last (at the end of the survey, preceding demographics). We pre-registered the following hypotheses: Participants in the concerns first case will be less accepting, less interested, and more concerned than those in the concerns last case and used independent samples t-tests to analyze mean differences across conditions.

## Results

### Participants

For the main sample (“Sample 1”), we recruited 1535 participants. Of those who began the survey, 1491 completed the study and 1435 participants correctly answered five or more of the comprehension checks (96.2%). See Table 1 for demographic information. In a second sample of 202 Prolific participants, 200 completed the full survey and 192 passed five or more of the comprehension checks correctly (96%). See Table 1 for demographic information.

**Table 1.** Demographics of Sample 1 and Sample 2.

|  | Sample 1<br>(n=1427) | Sample 2<br>(n=192) | Sample 2a<br>Concerns First<br>(n=95) | Sample 2b<br>Concerns Last<br>(n=97) |
| --- | --- | --- | --- | --- |
| **Age** |  |  |  |  |
| Mean (SD) | 45.8 (16.0) | 37.7 (12.2) | 36.5 (11.3) | 38.8 (12.9) |
| **Gender** |  |  |  |  |
| Female | 724 (50.7%) | 74 (38.1%) | 38 (40%) | 36 (37.1%) |
| Male | 656 (46.0%) | 110 (56.7%) | 51 (53.7%) | 59 (60.8%) |
| Other | 47 (3.3%) | 8 (4.1%) | 6 (6.3%) | 2 (2.1%) |
| **Race** |  |  |  |  |
| American Indian, Native American, Alaska Native | 16 (1.1%) | 7 (3.6%) | 3 (3.2%) | 4 (4.1%) |
| Asian | 91 (6.3%) | 22 (11.3%) | 9 (9.5%) | 13 (13.4%) |
| Black or African American | 189 (13.2%) | 18 (9.3%) | 7 (7.4%) | 11 (11.3%) |
| Native Hawaiian, Pacific Islander | 2 (0.1%) | 1 (0.5%) | 1 (1.1%) | 0 (0.0%) |
| Other | 20 (1.4%) | 2 (1.0%) | 2 (2.1%) | 0 (0.0%) |
| White | 1150 (80.1%) | 155 (79.9%) | 78 (82.1%) | 77 (79.4%) |
| **Ethnicity** |  |  |  |  |
| Non-Hispanic | 1346 (94.3%) | 178 (92.7%) | 89 (93.7%) | 89 (91.8%) |
| Hispanic or Latino | 81 (5.7%) | 14 (7.3%) | 6 (6.3%) | 8 (8.2%) |
| **Education level** |  |  |  |  |
| Less than Bachelor's | 642 (45%) | 94 (49.0%) | 41 (43.2%) | 53 (54.7%) |
| Bachelor's or higher | 785 (55%) | 98 (51.0%) | 54 (56.8%) | 44 (45.3%) |
| **Household income** |  |  |  |  |
| \$0–\$49,999 | 596 (41.8%) | 81 (42.2%) | 34 (35.8%) | 47 (48.5%) |
| ≥ \$50,000–109,999 | 553 (38.8%) | 77 (40.1%) | 44 (46.3%) | 33 (34.0%) |
| ≥ \$110,000 | 277 (19.4%) | 34 (17.7%) | 17 (17.9%) | 17 (17.5%) |
| **Parent of a child** |  |  |  |  |
| No | 707 (49.5%) | 122 (63.5%) | 61 (64.2%) | 61 (62.9%) |
| Yes | 728 (50.5%) | 70 (36.5%) | 34 (35.8%) | 36 (37.1%) |
| **Political Orientation** |  |  |  |  |
| Conservative | 361 (25.3%) | 43 (22.4%) | 24 (25.2%) | 19 (19.6%) |
| Liberal | 811 (56.9%) | 113 (58.9%) | 58 (61.1%) | 55 (56.7%) |
| Moderate | 254 (17.8%) | 36 (18.7%) | 13 (13.7%) | 23 (23.7%) |

*Note:* Sample 1 was recruited from Prolific with nationally representative quotas for gender, race/ethnicity, and age. Sample 2 was also recruited from Prolific but did not have a nationally representative quota for demographic characteristics. Participants randomly assigned to sample 2a were presented with the concerns first, while those assigned to sample 2b were presented with the concerns last (at the end of the survey). In both samples participants could select multiple races, therefore, percentages sum beyond 100%.

### Sample 1

#### Approval

As depicted in the first Likert plot of Figure 1A, 72% of the public either approved or strongly approved of people using polygenic embryo screening. Only 10% of the public either disapproved or strongly disapproved and 17% were ambivalent. These attitudes were similarly reflected in voting intentions with 77% of the public stating that they would vote to allow the use of PES, only 12% who would vote to disallow it, and 11% who would abstain from voting (Figure 1B). When gauging the public’s perception of PES’s risk-to-benefit ratio, 67% believed the benefits of allowing people to access PES outweigh the risks, 11% believed the risks outweigh the benefits, and 22% believed the risks and benefits to be equivalent (Figure 1C).

**Figure 1.**
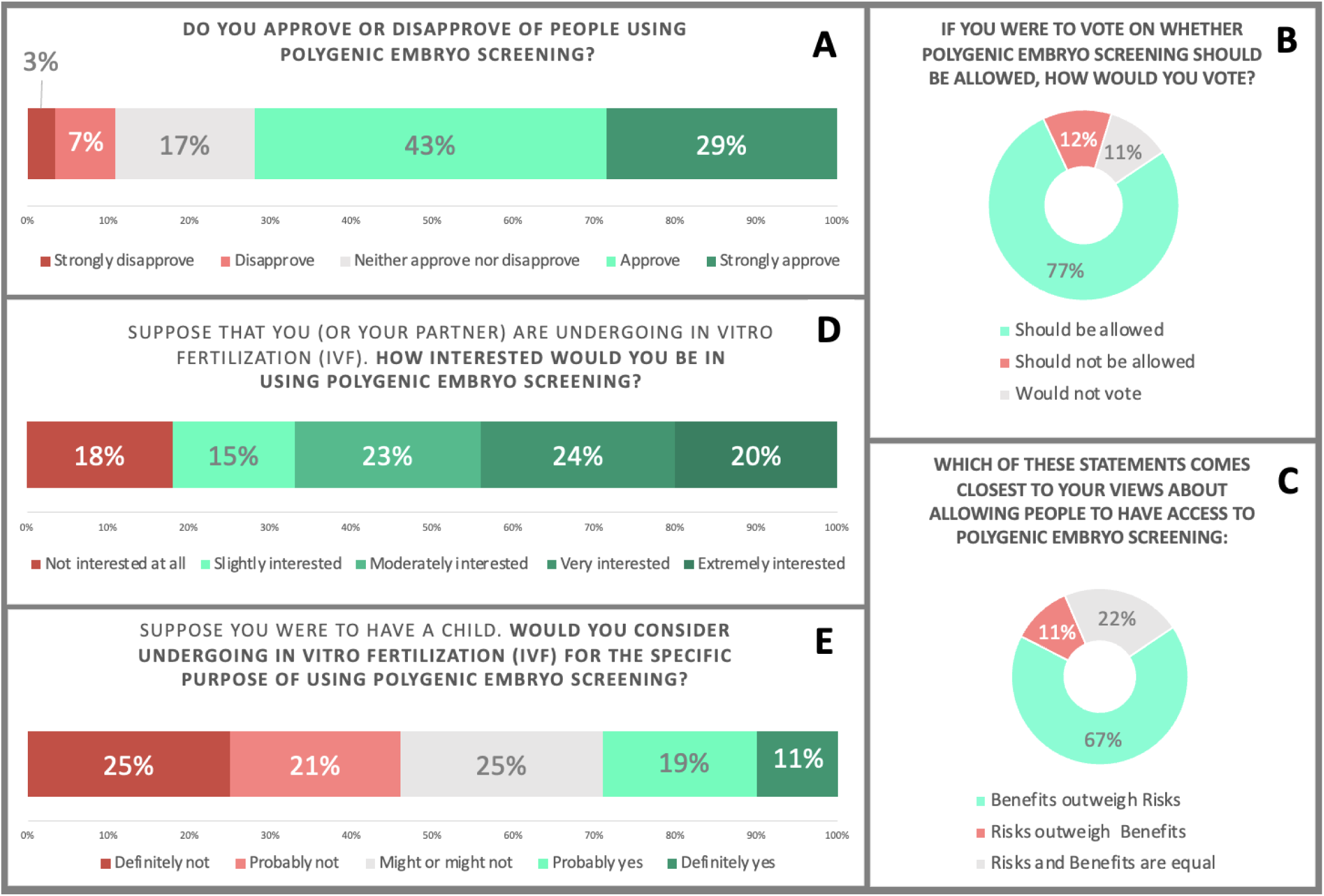
PES: Approval and Interest. *Note:* Sample responses for these items varied from N=1427-1428. Percentages are rounded to the nearest whole number, which, if summed according to the figures, can lead to percentage points being above or below 100%.

#### Interest

When participants were asked to gauge their interest in using PES under the circumstances of already undergoing IVF, 82% reported being at least slightly interested (Figure 1D). When asked whether participants would consider undergoing IVF for the specific purpose of getting access to PES, 30% reported they would “probably” or “definitely” consider using it, while 46% reported they would “probably” or “definitely” not consider using it, and 25% might or might not consider using it (Figure1E). Participants who answered that they were at least slightly interested in PES (n=1170) were also asked to report the maximum amount they would be willing to pay to access PES with the assumption that the cost of IVF was covered. Results ranged from $0 to $100,000 with the mean willingness to pay being $3240 and an interquartile range of $2600 (median= $1000, SD= $815).

#### Approval: Purpose x Outcome

Descriptive results of approval for different purposes (see below) across outcomes are depicted in Figure 2. Responses across the approval matrix of outcomes and purposes were coded numerically (approve=1, disapprove=-1, and no opinion=0)

**Figure 2.**
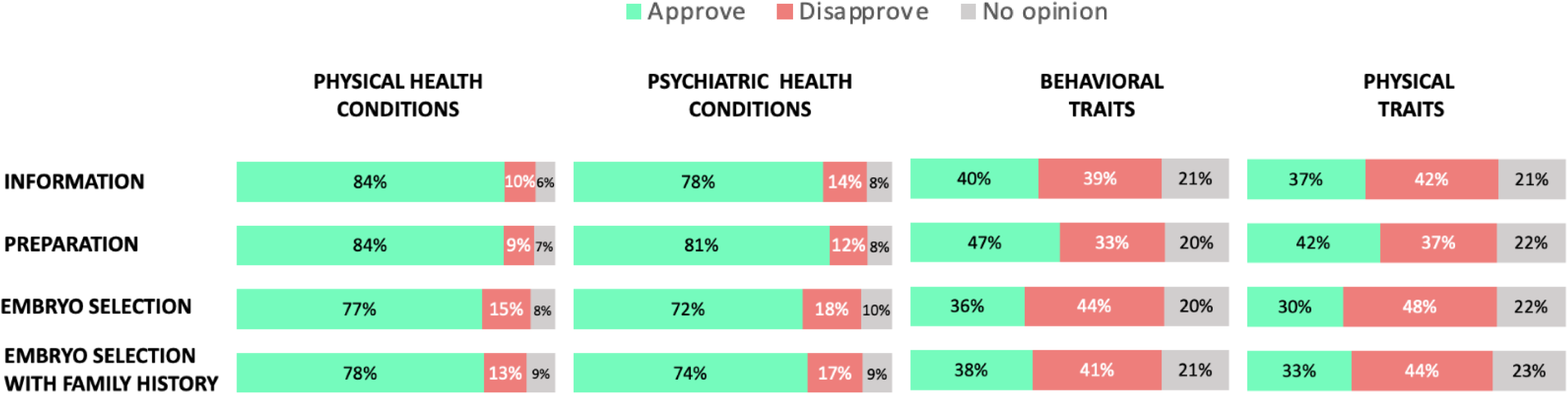
PES approval for varying purposes and outcomes. *Note:* Participants (n=1427) were asked to answer the following prompt: “ I **approve/disapprove of people using polygenic embryo screening for the following purposes based on the listed conditions and traits.** Purposes (embryo selection with family history, embryo selection, preparation, information) were presented to participants in the following way: **Information:** People using PES to know more about their future child. **Preparation**: People using PES to prepare (resources) for their future child. **Embryo Selection:** People using PES to select embryos to use with their preferred genetic chances. **Embryo Selection given family history:** People using PES to select embryos to use with their preferred genetic chances for traits or conditions that run in their family.

##### Purposes (information or selection)

Responses were summed across outcomes to test whether there were significant differences in approval between purposes: preparation (Mean (*M)*=1.63, Standard error of the mean (*SE*)=0.06), information (Mean (*M)* =1.36, (*SE)*=0.06), embryo selection based on family history (*M*=1.07, *SE*=0.07), and embryo selection (*M*=0.90, *SE*=0.07). Mauchly’s test of sphericity was significant, which required the use of Huynh-Feldt corrections. A repeated measures ANOVA demonstrated significant differences in approval across purposes *F*(2.34, 3336.67) = 110.49, *p*<.001, partial η^2^ =0.072. Post hoc tests using Bonferroni corrections revealed significant differences across all purposes *p*<.001.

##### Outcomes (traits and conditions)

Responses were summed across purposes to test whether there were differences in approval between outcomes: health conditions (*M*=2.75, *SE*=0.63), psychiatric conditions (*M*=2.44, *SE*=0.70), behavioral traits (*M*=0.60, *SE*=0.85), and physical traits (*M*=-.29, *SE*=0.83). Mauchly’s test of sphericity was significant, which required the use of Huynh-Feldt corrections. A repeated measures ANOVA demonstrated significant differences in approval across outcomes *F*(1.72, 2446.03) = 911.13, *p*<.001, partial η^2^ =0.39. Post hoc tests using Bonferroni corrections revealed significant differences across all outcomes *p*<.001.

#### Approval: screening conditions and traits

We asked participants to rate their approval for screening embryos on 12 conditions (physical and psychiatric; see left plot of Figure 3) and 12 traits (behavioral and physical; see right plot of Figure 3). Physical health conditions received the highest approval for screening (e.g., cancer (81%), heart disease (79%), and Alzheimer’s disease (77%)), with less than 15% of the public disapproving of screening for those conditions. Schizophrenia received the highest screening approval of the psychiatric health conditions, with 77% of the public approving, 9% being ambivalent, and only 14% disapproving.

**Figure 3.**
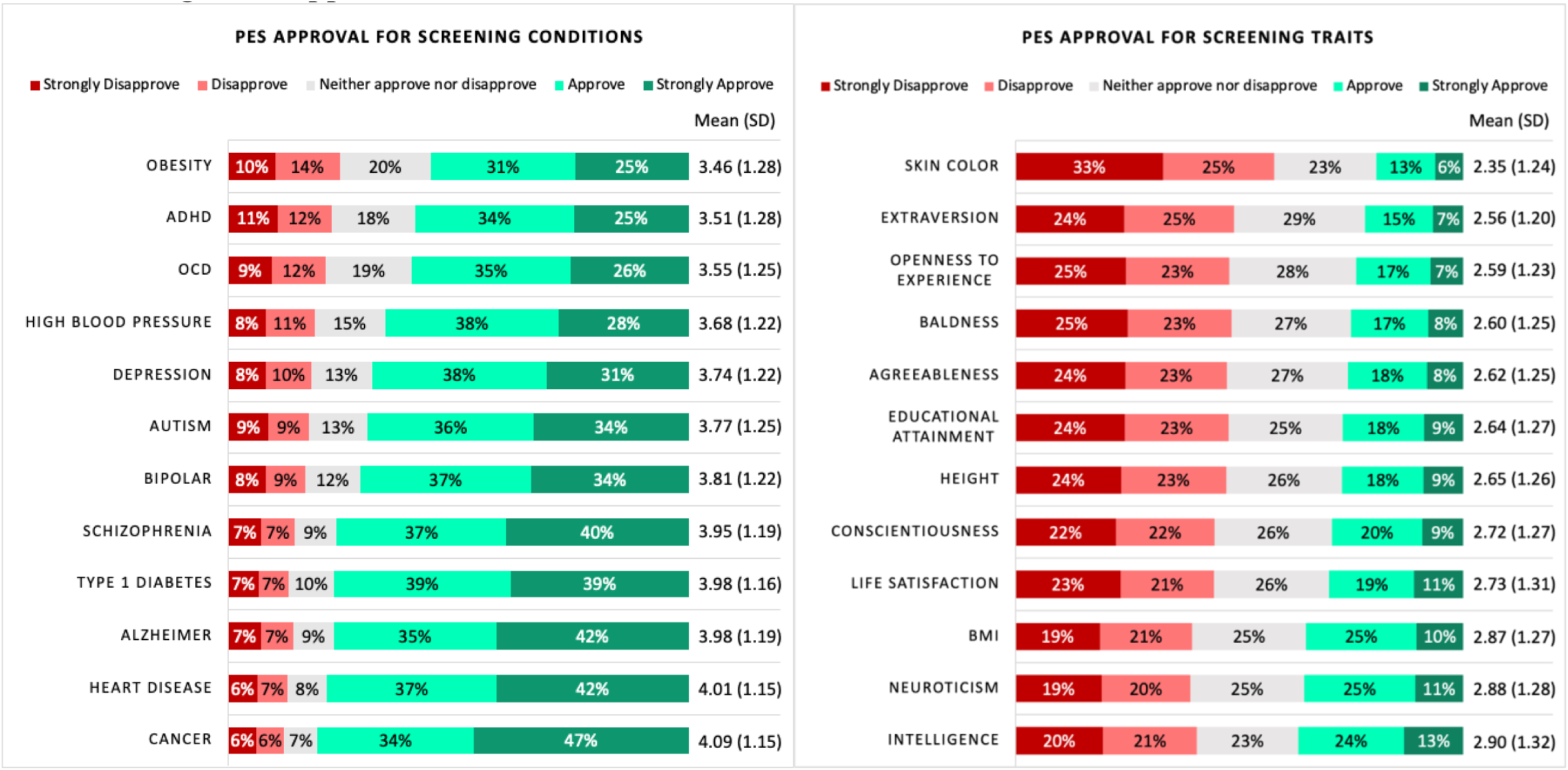
Approval for conditions and traits. *Note:* The left panel depicts approval ratings for conditions ranked by mean approval (in ascending order). The right panel depicts approval ratings for traits ranked by mean approval (in ascending order). Sample responses for these items were N=1427, except for depression and cancer N=1428. Percentages are rounded to the nearest whole number, which, if summed according to the figures, can lead to percentage points being above or below 100%.

The (physical) trait with the lowest approval for screening was skin color (58% disapproval), yet 42% of the public was either ambivalent (23%) or approved (19%) of screening for it. The highest approval was for screening the behavioral trait of intelligence, which 60% of the public either approved (37%) or was ambivalent (23%).

As pre-registered, we conducted a paired samples t-test to demonstrate that the mean approval rating (measured on a 5-point Likert scale) across the 12 traits (*M*=2.68, *SD*=1.10) was significantly lower than the mean approval rating across the 12 conditions (*M*=3.79, *SD*=1.08) *t*(1426)=-38.50, *p*<.001, *d*=-1.02.

#### PES: Potential Concerns

As demonstrated in Figure 4, when asked; “How concerned, if at all, are you with the following 13 potential concerns that have been raised about polygenic embryo screening (PES)?”, the proportion of participants who were at least “slightly concerned” was 68-91% for all 13 concerns. The proportion of participants who were “very” to “extremely” concerned was 50-55% for the following concerns: parents having false expectations about the future child, promoting eugenic thinking/practices, stigmatization of certain traits and conditions viewed as less desirable, treating embryos like a product by selecting them based on preferred genetic chances for conditions/traits, that it cannot be applied equally to all ethnicities, and the increased inequality due to its high costs. Overall, other than having false expectations, the public appeared less concerned about potential harm to themselves as patients, compared with harm to the future child or society.

**Figure 4.**
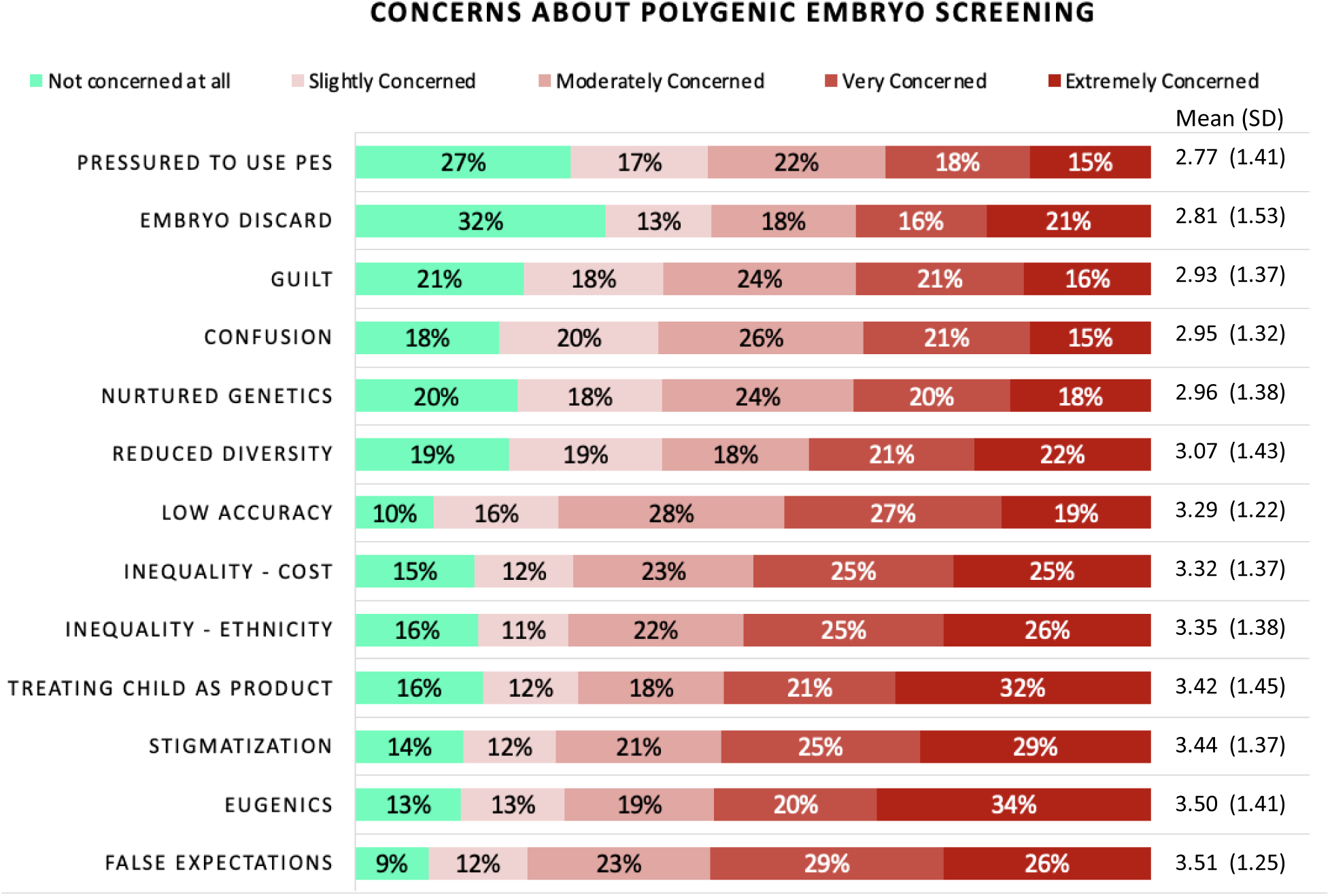
Potential concerns. *Note:* Participants were asked: “How concerned, if at all, are you with the following 13 potential concerns that have been raised about polygenic embryo screening (PES)?” and answered on a 5-point Likert scale. The 13 concerns presented in the figure are ranked by mean approval (in ascending order). Sample size varied from N=1420-1424 across concern items.

##### An experiment comparing concerns first vs last

In sample 2 (n=192), experimentally randomizing the presentation of potential concerns significantly decreased general approval between those who were presented with concerns last (*M*=3.90, *SD*= 0.94), and those who were presented with concerns first (*M*=3.36, *SD*=0.87), *t*(190) = 4.1, *p* <.001; *d*=.59 (See Figure.6). General approval decreased from 71% to 43%, neither approving nor disapproving increased from 19% to 43%, and general disapproval increased from 10% to 14%. Participants’ interest also significantly decreased between those who were presented with concerns last (*M*=3.53, *SD*=1.47) and those who were presented with concerns first (*M*=2.78, *SD*=1.29), *t*(190) = 3.75, *p* <.001; *d*=.54 (See Figure 6.). No significant differences were detected on a concern index (averaging across the thirteen concerns) between those who were presented with concerns last (*M*=3.11, *SD*=0.77) and first (*M*=2.97, *SD*=0.80), *t*(190) = 1.24, *p*=.22; *d*=.18. All other t-test results comparing concerns first vs last results are reported in the supplemental material.

**Figure 6.**
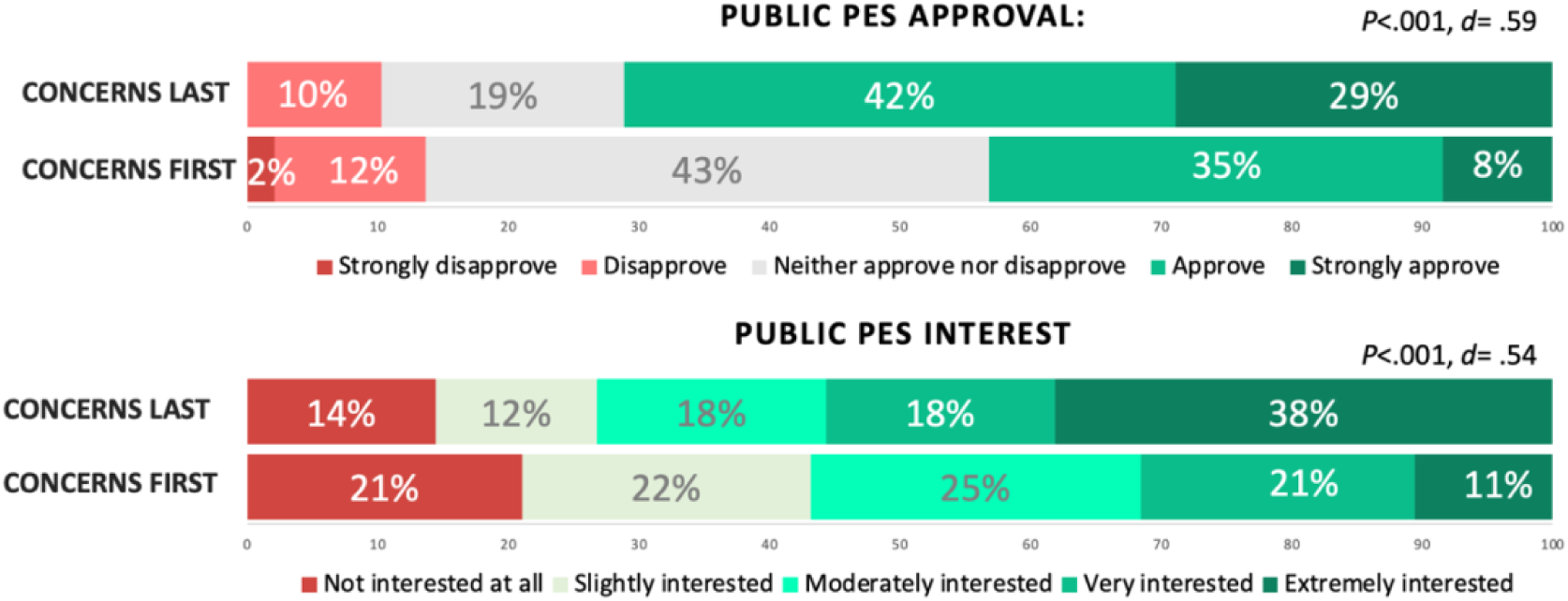
Randomizing PES concerns (first vs. last): approval and interest in PES. *Note:* The top Likert plot depicts the (%) distribution of PES approval for concerns first (sample 2a) and concerns last (sample 2b). The bottom Likert plot depicts the (%) distribution of PES interest (assuming they were undergoing IVF) for concerns first (sample 2a) and concerns last (sample 2b)

## Discussion

Our present findings demonstrate that within the US public, 72% approve of using PES. When specifying that the purpose of PES is for embryo selection, 77% approve of using PES for screening physical health conditions and 72% for screening psychiatric conditions. In contrast, there is minority approval for embryo selection based on behavioral traits (36%) and physical traits (30%). When specifying that the purpose of PES is for parents to prepare for their future child, approval increases to 84% for physical health conditions, 81% for psychiatric conditions, 47% for behavioral traits, and 42% for physical traits. The high level of public approval and interest for PES raises concerns over the lack of regulation and guidance on what type of polygenic embryo health conditions or traits should be screened and under what conditions.

Upon being informed of potential concerns, 54%-55% of the public report being “very” to “extremely” concerned about PES promoting eugenic practices or leading parents to have false expectations about their future child. Further investigation into the public’s conceptions of eugenics is warranted considering that this concern appears to be at odds with the high approval of screening embryos for health conditions for purposes of selection. ^37^.

In a second sample, we experimentally investigated the effect of presenting the list of potential concerns at the start of the survey (vs. at the end) on the public’s attitudes towards PES. Notably, we found that when concerns were presented at the start of the survey, general approval decreased by 28%, while uncertainty increased by 24%, but disapproval only increased by 4%. These findings suggest that while the public shares concerns related to PES, as raised by REIs and IVF patients, those concerns aren’t intuitively salient to the public, since their attitudes changed significantly when presented with the concerns. The decrease in approval could also be due to social desirability bias (i.e., responding in a manner thought to be viewed favorably) ensuing from being primed with a list of concerns. Additionally, because the effect only raised the level of concerns minimally and mostly raised doubt as opposed to disapproval, it further suggests that the public does not find the concerns sufficient to disapprove of PES.

These findings highlight the urgent need for further research to comprehensively inform the public, IVF patients, REIs, and genetic counselors on the potential benefits, limitations, and concerns that surround the use of PES. Given the divide in approval between the public and IVF patients on the one hand and healthcare professionals on the other^22,25,34–36,38^ we recommend that stakeholder groups, particularly policymakers, working groups from professional societies, and clinicians, consider the basis of this divide. Finally, approval was significantly different between using PES for health conditions versus traits. One issue to be considered by policymakers, including professional medical organizations, is whether to move ahead with regulating the use of PES for non-medical traits (physical and behavioral) which would align with our findings that only a minority of the public approves of screening for traits. PES is already commercially available, and physicians and patients need guidance about whether and how to use these technologies. Professional medical societies in the United States are in the best position to publish guidelines that can quickly help set a standard of care regarding the use of PES in the IVF context.

## Supplemental Materials

https://docs.google.com/document/d/1fAGLS8bmHC9FX-Ki0Suey3Y1065oGrrq2wydMKTYmI0/edit

## Data Availability

All data produced in the present study are available upon reasonable request to the authors

https://docs.google.com/document/d/1fAGLS8bmHC9FX-Ki0Suey3Y1065oGrrq2wydMKTYmI0/edit

